# 3D Dental Similarity Quantification in Forensic Odontology Identification

**DOI:** 10.1101/2025.03.27.25324745

**Authors:** Anika Kofod Petersen, Andrew Forgie, Palle Villesen, Line Staun Larsen

**Affiliations:** Department of Forensic Medicine, Aarhus University, Denmark; School of Medicine, Dentistry and Nursing, University of Glasgow, Scotland; Bioinformatics Research Centre, Aarhus University, Denmark; Department of Clinical Medicine, Aarhus University, Denmark; Department of Dentistry and Oral Health, Aarhus University, Denmark

**Keywords:** Keywords Forensic Odontology Identification, Disaster Victim Identification, Keypoint Detection, Keypoint Description, 3D Dental Comparison, Biometric Identification

## Abstract

Forensic odontology identification largely depends on comparing dental work like fillings and crowns with the dental records of potential victims. This process can be challenging, especially regarding victims with minimal or no dental work. Alternatively, 3D tooth morphology can be used for identification by automated dental surface similarity scoring. However, high-resolution 3D intraoral photo scans contain hundreds of thousands of datapoints from each individual jaw, making database searches difficult and slow.

Here, we reduce full 3D scans to keypoints, which are small points located in areas of high curvature on tooth surfaces. We use Difference of Curvature (DoC) for robust keypoint detection and evaluate different keypoint representation methods to distinguish between scans of the same individual and scans of different individuals, assigning them a similarity score.

The results demonstrate that combining DoC with the Signature of Histograms of OrienTations (SHOT) representation method effectively separates matches from mismatches. This indicates the potential for automatic scoring of dental surface similarity. This can be valuable for forensic odontology identification, especially in cases where traditional methods are limited by the lack of dental work.

## 1. Introduction

According to Interpol [1, 2], forensic odontology identification is one of the three primary identifiers for identifying deceased victims during Disaster Victim Identification (DVI). In the wake of a disaster with human casualties, forensic odontologists are tasked with assessing the similarity of the dentitions described in *ante mortem* (AM) dental records of the known missing individuals with post mortem (PM) examinations of the dentitions of the deceased. The outcome range of the comparisons is *‘Identification with absolute certainty’, ‘Identification probable’, ‘Identification possible’ to ‘Identity excluded’ or ‘Insufficient evidence’* [2]. Each conclusion is thus an assessment of individualization based on the forensic odontologist’s experience and the human dentition including the amount of AM and PM data available holding specific characteristics [3]. The uniqueness of the human dentition has been widely discussed in the literature [3-6], and only recently proven theoretically on a molecular level, thus questioning whether identification based on dentitions is plausible [4]. Nevertheless, the human dentition is continuously used worldwide as a part of forensic science both for identification cases and for finding the source of bitemarks [2, 4, 7, 8]. Interpol’s DVI guide highlights the importance of dental work, e.g. dental fillings and prosthetics, for the forensic odontologist to enable the conclusion *identification with absolute certainty*, alongside dental morphology [2]. This addition of dental work, on top of dental variation, aids in differentiation between, and thus to some part undermines the importance of dental uniqueness [2, 3, 5]. But in the case of individuals with little or no dental work, identification is often very difficult to conclude with absolute certainty for the forensic odontologist. However, indication of identification is still possible using the individuals’ tooth morphology only. The anatomical traits of teeth, especially detailed crown morphology, which at present are relatively unexploited within common DVI, could prove crucial in the identification process [2, 9]. A worry with classic manual tooth morphology comparison in forensic odontology identification is the involvement of a certain amount of qualitative subjective assessment, especially considering the sensitivity to perspective in 2D imaging [10, 11]. An objective quantitative measure of dental surface similarity would be beneficial for the continuing credibility of the identification process [2, 7, 8, 12].

In recent years, the use of 3D intraoral photo scans in dentistry has increased [13, 14], thus increasing the number of AM dental records containing digital 3D models of clinically visible tooth morphology. These digital models could, in a disaster situation, be collected to form an AM 3D dental database to be used in the identification process. To automate parts of this identification process, a PM 3D dental scan could be used to search an AM dental database and return the most likely matches, giving a quantitative similarity score [14-18]. Such a search method should be able to handle adverse disaster conditions, tooth loss and natural tooth movement with time, wherefore conventional 3D matching (superimposition) does not suffice, as such methods are unable to handle the difference between intrapersonal and interpersonal discrepancies [12, 15-17, 19, 20].

Miki and colleagues [16] showed promising results, exploiting the morphology of the molar occlusal surfaces to perform AM-PM matching, indicating signature dental morphologies unique enough to perform robust AM-PM matching. However, they reduced the 3D models to 2D images before performing AM-PM comparisons, thus losing information that could impact the results of AM-PM matching, while also limiting the scope of comparison to molars only [16]. 3D dental surface meshes are models of surfaces that consists of hundreds of thousands of vertices (points in space) connected by edges (lines between vertices) [21]. When using 3D dental surface meshes, comparison of all vertices between meshes is suboptimal since many vertices in smooth areas hold very little informative information (e.g. dental buccal and lingual surfaces) and the comparison will be computationally heavy, and more noise may be introduced [15, 17, 22]. A simple strategy to optimise mesh comparison is to focus on keypoints: small areas of great curvature that encodes essential information (e.g., dental cusp tips and ridges).

This paper explores the robustness of keypoint detection and the accuracy of dental comparisons when using keypoints covering the dental curvature landscape rather than full 3D scans in an AM-PM matching scenario. The specific aim was to be able to automatically score 3D dental comparisons in order to find possible matches of identity within a database.

## 2. Methods

Two datasets of different size were used in this study (Figure 1). The smaller dataset of 6 dental meshes from 3 different individuals was initially copied and some of the surface meshes were added noise (augmentation). This dataset was used to estimate optimal resolution of the 3D scans and to estimate the optimal distance threshold for vertices to be included for keypoint description (support radius). The larger dataset of 60 dental meshes from 30 individuals was augmented followed by keypoint detection, keypoint description and keypoint correspondence for performance evaluation.

**Figure 1:**
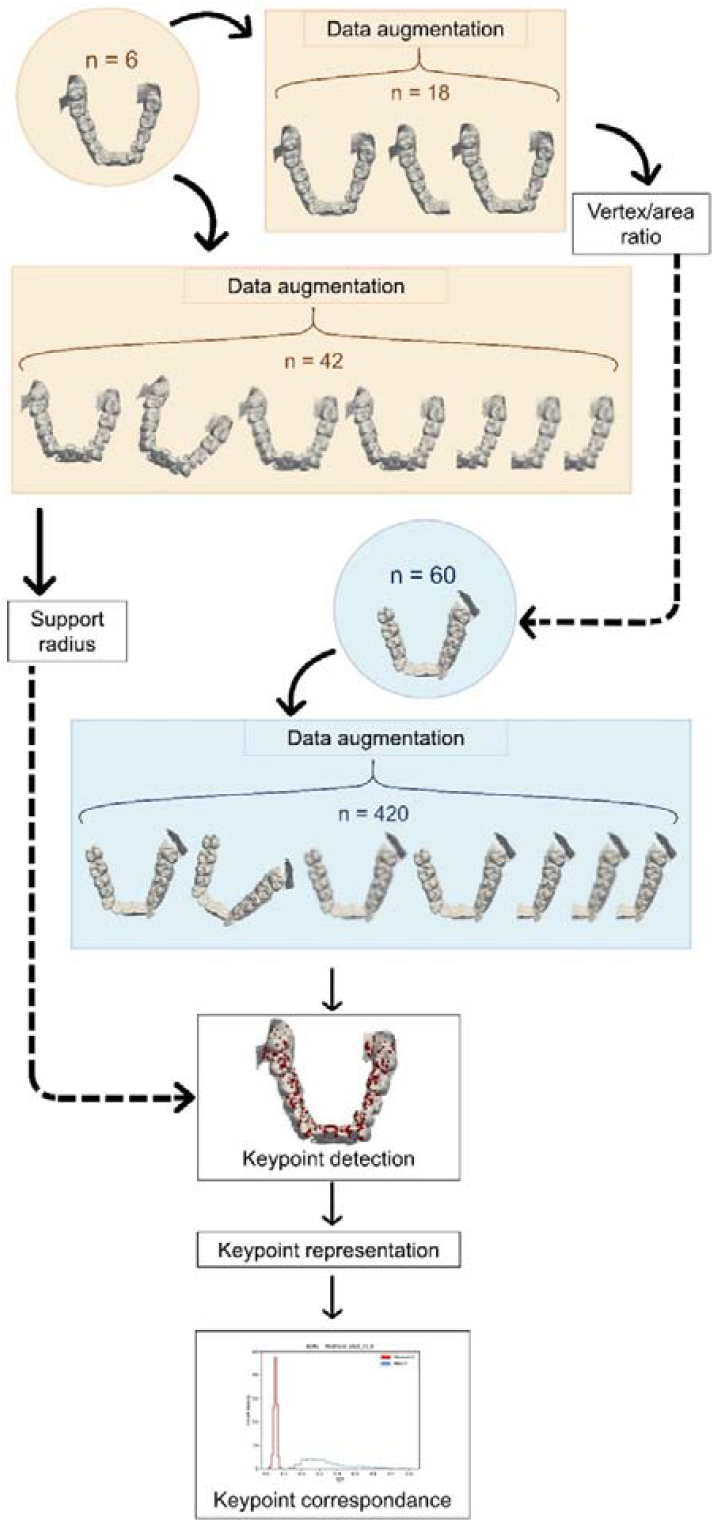
Method overview. Two datasets were used for this study. The smaller dataset (n=6) were subjected to augmentation and used to determine the 3D resolution parameter for keypoint detection and the support radius parameter for keypoint representation. The larger dataset (n=60) was subjected to augmentation, followed by keypoint detection. The detected keypoints were subject to description followed by correspondence, where keypoints originating from different dentitions are compared. Keypoint correspondence was used to evaluate if the keypoints originated from the same dental scan or different dental scans.

Keypoint detection consists of locating curvature positions in the dental 3D mesh. To simply compare the X, Y, Z-coordinates of keypoints will not allow for matching unless the AM and PM scans have identical placement and rotation, which will rarely be the case between scans. Instead, a representation is a way of describing an object or structure using numbers, such as the position in space (e.g. x, y, z) and/or details about the local shape around the keypoint. Choosing the right representation before comparing keypoints is important because it affects how accurately and efficiently keypoints can be matched.

All data processing was performed using custom Python code, which can be found at https://github.com/AnikaKofodPetersen/Dental-Similarity-Quantification.

### 2.1 Data

This study was registered with the Data Protection Unit at Aarhus University, Denmark (file number 2016-051-000001, serial number 2534; file number 20220367531, serial number 3155). It was conducted in full accordance with the World Medical Association Declaration of Helsinki and complied with the European Union General Data Protection Regulation legislation. According to the assessment by the chairmanship of The Danish National Committee on Health Research Ethics (NVK), the study and its protocol are deemed exempt from notification (case number: 2400741). The two datasets shown in Figure 1 have previously been described by Kofod Petersen and colleagues [18]. Briefly, the dataset used for parameter optimization consisted of intraoral 3D photo scans in the form of dental surface meshes of 6 dental arches from 3 individuals [18] (Figure 1). The dataset for the analysis of keypoint detection and keypoint description consisted of intraoral 3D photo scans in the form of dental surface meshes of 60 dental arches from 30 healthy individuals who volunteered and gave informed written consent to participate in the study [18] (Figure 1).

For testing the reproducibility of keypoint detection, all dental meshes were augmented twice. One copy was subjected to decimation, where vertices were removed from the dental mesh to lower the mesh resolution. This copy was decimated to keep 95% of all vertices. The same copy was then subjected to remeshing, where the remaining vertices were re-placed on the mesh surface to ensure even vertex coverage and accurate modelling of the original mesh surface. This copy was hereafter referred to as the remeshed copy. The other copy was cut through the centre of mass, in the xz-direction, followed by decimation to 95% of all vertices and remeshed. This copy was hereafter referred to as the partial copy. The dataset ended up with a total size of 18 mesh surfaces, describing 6 different dental arches (Figure 1).

To simulate multiple intraoral 3D scans from the same individual, while including the estimated systematic differences that could occur by using different intraoral scanners, we chose different strategies. Each dental surface mesh was augmented into 3 copies. The three copies were then either rotated and remeshed or had some noise added. One copy was rotated 17 degrees around the z-axis, decimated to 80% of the original vertices, remeshed, and moved 100 mm out of centre.

The two other copies were distorted using Gaussian noise, with standard deviation 0.1*the mean edge length (MEL), and 0.3*MEL, respectively, as used by Buch *et. al* [23] and Zhang *et. al* [15].

We also did the same for partial meshes, thus simulating matching partial dentitions. Here, we cut the original mesh through the centre of mass with a plane in the xz-direction, preserving the teeth placed distally, since these teeth would be less likely to be impacted by disaster conditions. This cutting was followed by decimation to keep 85% of the original vertices and remeshing to ensure an even coverage of vertices and finally moved 100 mm out of centre. We further distorted this partial copy into two more copies with Gaussian noise added (again standard deviation of 0.1 and 0.3* MEL).

In total we augmented each mesh into 6 distorted copies, resulting in a total of 420 meshes from the original 60. To avoid adding bias when performing dental comparisons, comparison is only performed on non-identical dental scans originating from the same jaw (maxilla or mandible), where a maximum of one of the compared dental scans have been added gaussian noise. This gives 1.800 matches to be differentiated from 57.420 mismatches.

### 2.2 Keypoint detection using Difference of Curvature (DoC)

Instead of focussing on the entire dental surface mesh, points of interest, also known as keypoints, was the focus of this analysis. Difference of Gaussian (DoG) is a method frequently used for edge detection that has previously been used for keypoint selection in 2D images [24, 25]. In the case of 3D meshes, a similar method called Difference of Curvature (DoC), has proven useful [26].

For this study, DoC was initialized by decimating the mesh to a fixed 3D resolution (measured as vertices per surface area), followed by smoothing using six Gaussian filters, creating five scales. To decide on an appropriate 3D resolution, the minimal distance between keypoints found on the original scan, a remeshed copy and a partial copy was measured. This ensured that keypoints were placed at the same positions of interest.

Inspired by Tang *et. al* the maximum and minimum principal curvature measures were used to find keypoints using the difference of principal curvatures on the five scales [26]. To find the maximum and minimum principal curvatures, the mean curvature and the Gaussian curvature for the dental surface meshes were calculated using *The Visualization Toolkit* (VTK) [27]. Since the mean and Gaussian curvatures for the boundaries of the dental meshes were unstable, the curvature values for the boundaries were set as follows:

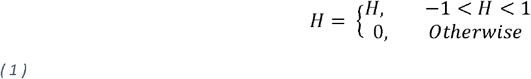

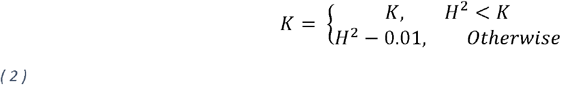

Where H is the mean curvature and K is the Gaussian curvature. This ensures that complex numbers are not encountered when calculating the principal curvatures. Since the mesh boundaries were not considered for keypoint detection, these definitions did not have an impact on the final keypoint detection but merely eased the calculation of the principal curvatures. The principal curvatures were then found for each vertex as follows:

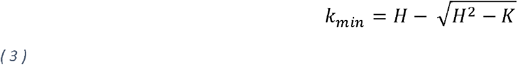

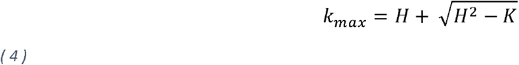

Where k_min_ is the minimal principal curvature, k_max_ is the maximum principal curvature. By comparing principal curvatures of adjacent scales, extrema of curvature changes were identified, indicating vertices that could be used as keypoints [26].

As suggested by Lowe *et. al*, adding a contrast threshold could make the approach more robust [25]. For a vertex to be classified as an extremum, and thus a potential keypoint, the difference of one of the principal curvatures within a one-ringed neighbourhood must be more than 0.00001. Furthermore, Lowe *et. al* describes how DoG is unstable along the boundary [25]. Therefore, as mentioned earlier, if a possible extremum vertex is a boundary vertex, it cannot be considered a keypoint. This approach allows for detecting keypoints that are placed on positions with most difference in curvature between scales as seen in Figure 2.

**Figure 2:**
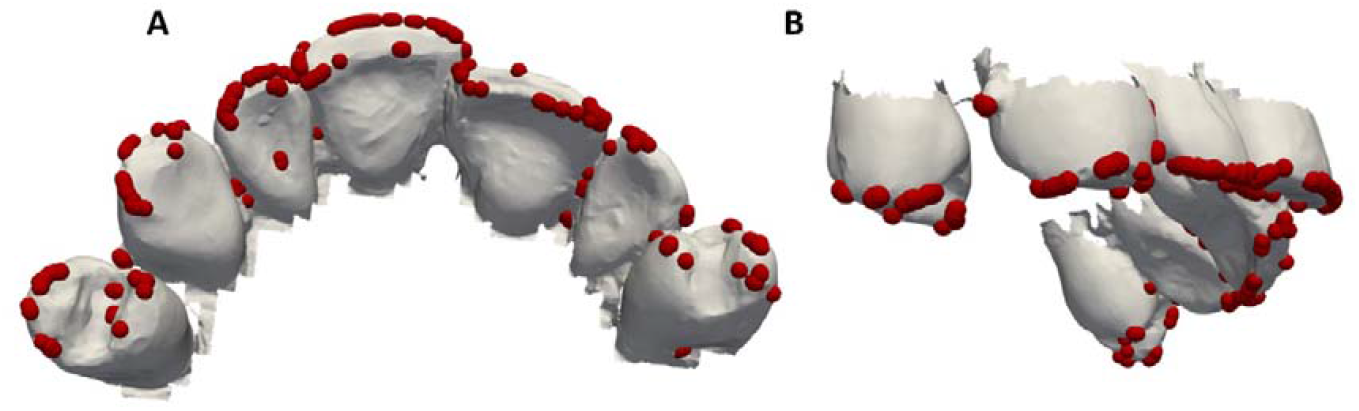
Keypoints. Keypoints (red spheres) found using DoC with 3D resolution of 40 on maxillary dentition after soft tissue removal using grid cutting [18]. Shown from A) occlusal view B) right side view.

### 2.3 Keypoint representation

To simply compare the X, Y, Z-coordinates of keypoints will not allow for matching unless the AM and PM scans have identical placement and rotation. To compare dental surface meshes using keypoints, one must therefore account for the slight differences in placement and rotation of the dental arch that can happen during scanning, and the slight natural drifting of teeth towards the centre line that happens over time [21]. To overcome this, the relation between a keypoint and the surrounding vertices, such as connectivity and angles can be considered; such relationships are referred to as keypoint representations. For dental matching, the keypoints must be represented in an invariant manner, that allows for differentiation between intraoral 3D photo scans from the same individual (matches) opposed to scans from different individuals (mismatches) [25, 28]. Buch *et. al* experienced significant performance differences when trying to find the best representation method to distinguish 3D shapes [23]. One conclusion from Buch *et. al* was that the best representation method might differ between datasets [23]. Four representation methods performed best overall in their analysis, namely Unique Shape Context (USC) [29], Signature of Histograms of OrienTations (SHOT) [30], Equivalent Circumference Surface Angle Descriptors (ECSAD) [31], and Rotational Projection Statistics (RoPS) [28]. In Addition, Signed Feature Histograms (SFH) has been suggested as a representation method specifically developed for dental surface meshes by Zhang *et. al* [15]. This study tests all five representation methods, and combinations hereof, to find a representation that allows for separation of matches and mismatches.

### 2.4 Comparison of scans

To determine which representation method to use, all five methods and a range of combinations were tested as suggested by Buch *et. al* [23]. Dental arches were subject to keypoint detection, with a 3D resolution of 40 vertices pr. mm^2^, followed by keypoint description/representation by the stated representation methods. Representations of a keypoint can be thought of as a descriptive vector, just like the Euclidean coordinates (x, y, z) is a descriptive vector for a point in space. Coordinates that are similar lie close together and will have a short distance between them. The same goes for the representations, even though the descriptive vector is longer than 3 coordinates. Therefore, the relative distance between keypoint representations across dental scans is a measure of how similar the keypoints are, and consequently how similar the two scans are. To estimate how similar two scans are, we must figure out how many keypoints of one scan have a matching keypoint on the other scan.

To do so, the representations of keypoints of each dental arch were compared by calculating the Euclidean distances (L2 distances) from one keypoint on one dental surface mesh to all keypoints in another dental surface mesh. These L2 distances are then sorted, and the ratio between the shortest and the second shortest distance is calculated. This gives one L2 ratio for each keypoint, as presented by Lowe *et. al* [25]. This was done for all 3D scans. Upon investigating the resulting L2 ratio distributions of the keypoint representation comparisons, the main difference between L2 ratios from matches of identity and mismatches of identity was assessed to be the Interquartile Range (IQR) of the L2 ratios of the comparisons (Figure 3). Each line in Figure 3 shows the cumulative histogram of density of L2 ratios for a comparison between two scans. The lines are coloured according to their classification. For each comparison, the IQR shows the range within which the middle 50% of the L2 ratios for that comparison fall. The IQRs of comparisons generally seems to be larger for matches than for mismatches, since the red lines have a later, and more drastic, rise in the cumulative histogram of densities (Figure 3).

**Figure 3:**
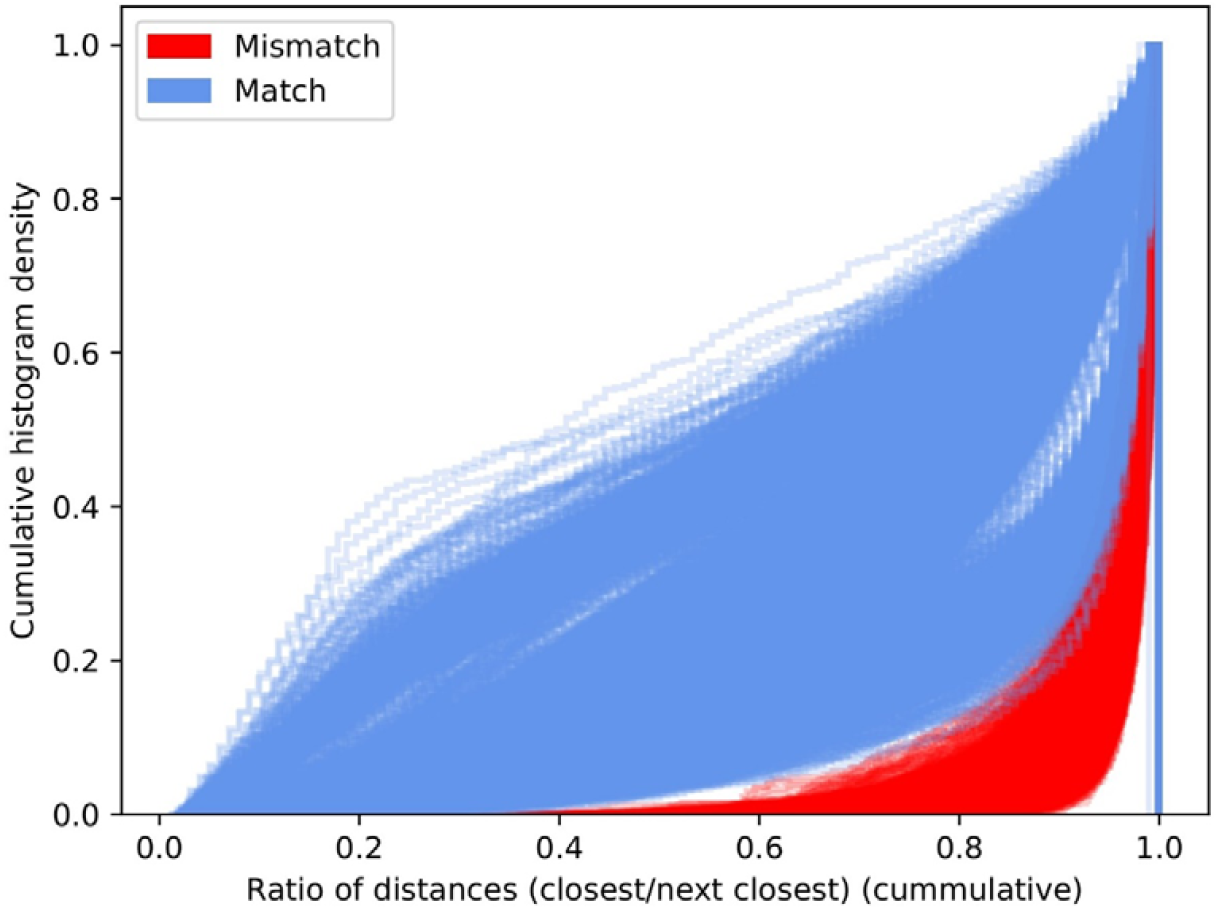
L2 ratio distributions. Cumulative distribution of L2 ratios for dental comparisons using SHOT representation, colored by the classification of the jaws being matched (matches/mismatches)

Since the aim of the study is to score the dental comparisons to perform an ordering of the dental arches according to a most likely match, the IQR was decided upon as a scoring system for the dental comparisons.

## 3 Results

### 3.1 3D resolution

To ensure robustness of keypoint detection, we tested what 3D resolution (vertices pr mm^2^) minimised variation of keypoint placement using the original scans, the partial copies and the remeshed copies of the 6 dental surface meshes described in section *2.1.* Since the remeshed copy and the partial copy are in fact copies of the original mesh, the positions of the dental meshes are the same, thus the Euclidean distance between the keypoints of the two copies and the closest keypoint of the original mesh estimates how robust the keypoint detection technique is. As seen by Figure 4, higher resolution ensures shorter distance between the keypoints of the original mesh and the keypoints of the copies. We chose 40 vertices pr mm^2^ since we found diminishing improvement of keypoints < 1 mm from the closest keypoint of the original mesh when comparing the two copies.

**Figure 4:**
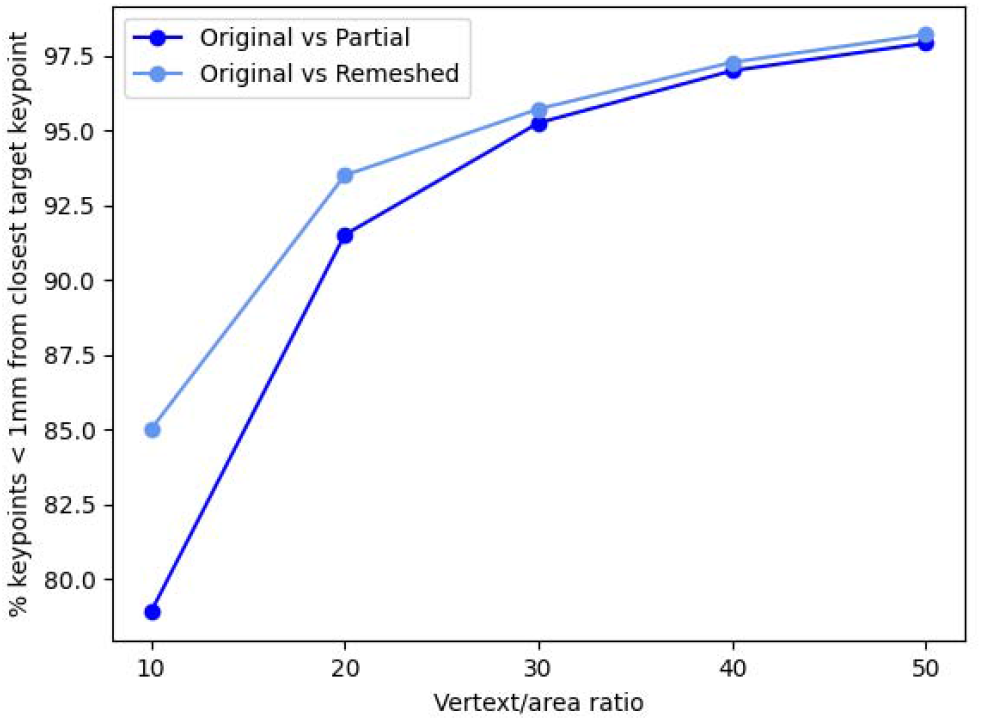
Keypoint detection robustness with increasing 3D resolution. Scree plot of percent of keypoints < 1 mm from the target keypoint when comparing the original mesh and its remeshed and partial copy when increasing the 3D resolution.

### 3.2 Support radius

The support radius describes the amount of surface area to include when describing a keypoint. It is important that this area covers enough surface for curvatures to show differentiation, but it is also important to limit the radius to make keypoints less dependent on tooth position. Therefore, the support radius should be between 1 and 3 mm. An in-house preliminary assessment showed promising results using SHOT representation to describe keypoints, which is why SHOT was used for keypoint representation in the augmented smaller dataset to set the final support radius. Increasing support radii were tested and the overlap of score used to differentiate between matches of identity and mismatches of identity (IQR) was tracked. Figure 5 shows that at 2 mm there is a margin between the IQR scores of matches and mismatches, while the margin decreases at 2.5 mm. Therefore, the support radius was set to 2 mm.

**Figure 5:**
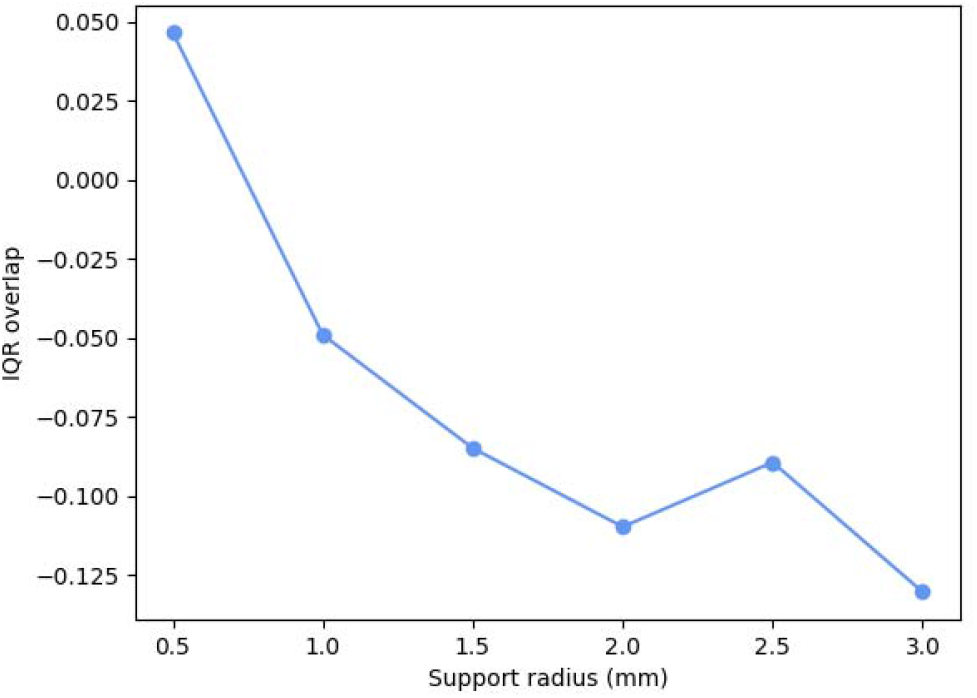
IQR overlap of the SHOT representation method with increasing support. Due to the decrease of margin at 2.5 mm, 2 mm was chosen as support radius.

### 3.3 Different representations

The augmented dental scans from the large dataset (Figure 1) were subject to keypoint detection followed by keypoint representation by RoPS, ECSAD, USC, SFH and SHOT (Figure 6).

**Figure 6:**
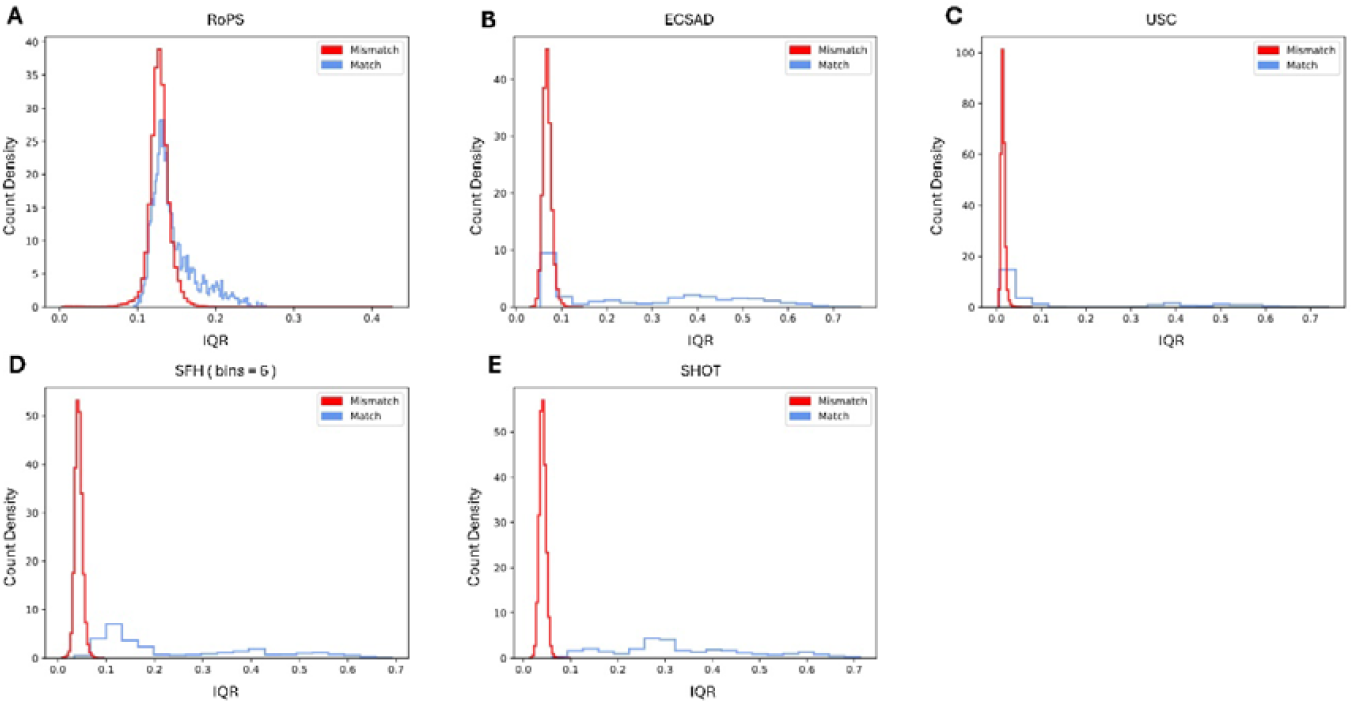
IQRs of the L2 ratios for each dental scan comparison. Shown as density histograms. Representation is made using support radius = 2 mm and representation method A) RoPS, B) ECSAD, C) USC, D) SFH, bins = 6, E) SHOT.

When two dental scans are compared, each method gives the comparison a single IQR score. This score is the quantitative measure of how well the dental scans are matching, using each representation method. For a representation method to be considered suitable for dental comparison, it must be able to distinguish between dental scans originating from the same individual (match) and dental scans originating from different individuals (mismatch). To assess whether a representation method has this capacity, the IQR scores are clustered according to their true label (match or mismatch). If a representation method is perfect at scoring matches and mismatches differently, the IQR score distributions for matches and mismatches will have no overlap. Therefore, the performance of a representation method can be found by assessing the part/share of overlap of the matching and the mismatching distribution.

As seen on Figure 6, RoPS is not able to separate matches and mismatches. ECSAD and USC are in some cases able to distinguish between matches and mismatches, but due to a significant overlap, these representation methods are not suitable for dental comparison. Both SFH and SHOT are able to distinguish between matches and mismatches in most cases, with the overlap of IQR using SHOT being the smallest.

Since SFH has not been thoroughly tested in the literature, three bin sizes were tested in this study (Figure 7). The increase in SFH bin size did not substantially change the IQR overlap. Therefore, SHOT is considered the most appropriate single representation method for dental comparison.

**Figure 7:**
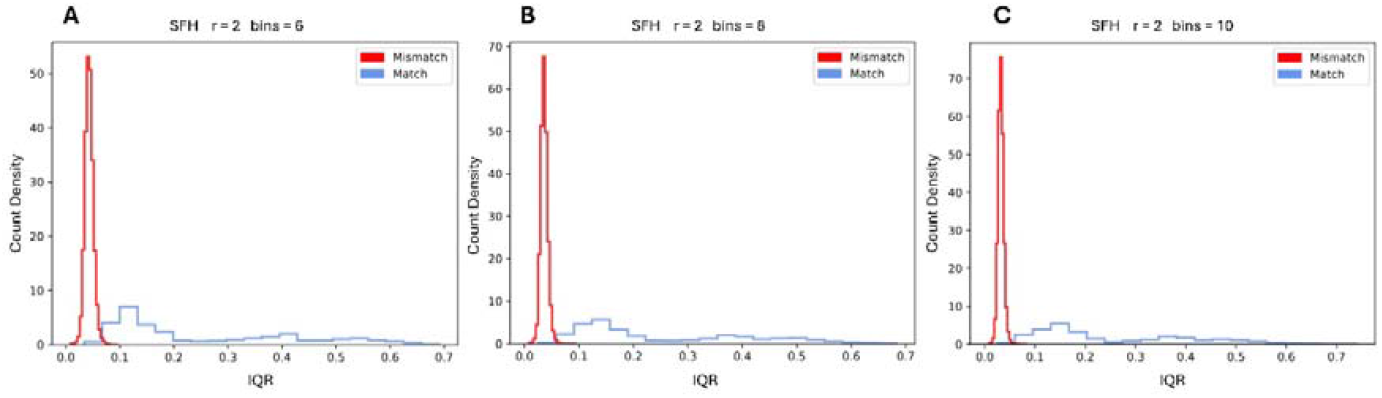
IQR scores of the L2 ratios for each dental scan comparison using SFH. Shown as density histograms. Representation is made using support radius = 2 mm and representation method A) SFH, bins = 6, B) SFH, bins = 8, C) SFH, bins = 10.

As suggested by Buch *et. al* [23], we concatenated representations, in hope of seeing a larger margin separating matches and mismatches than when using SHOT representation alone (Figure 8). None of the combinations showed separation without an overlap and the single method, SHOT, remained the representation method of choice for dental comparison.

**Figure 8:**
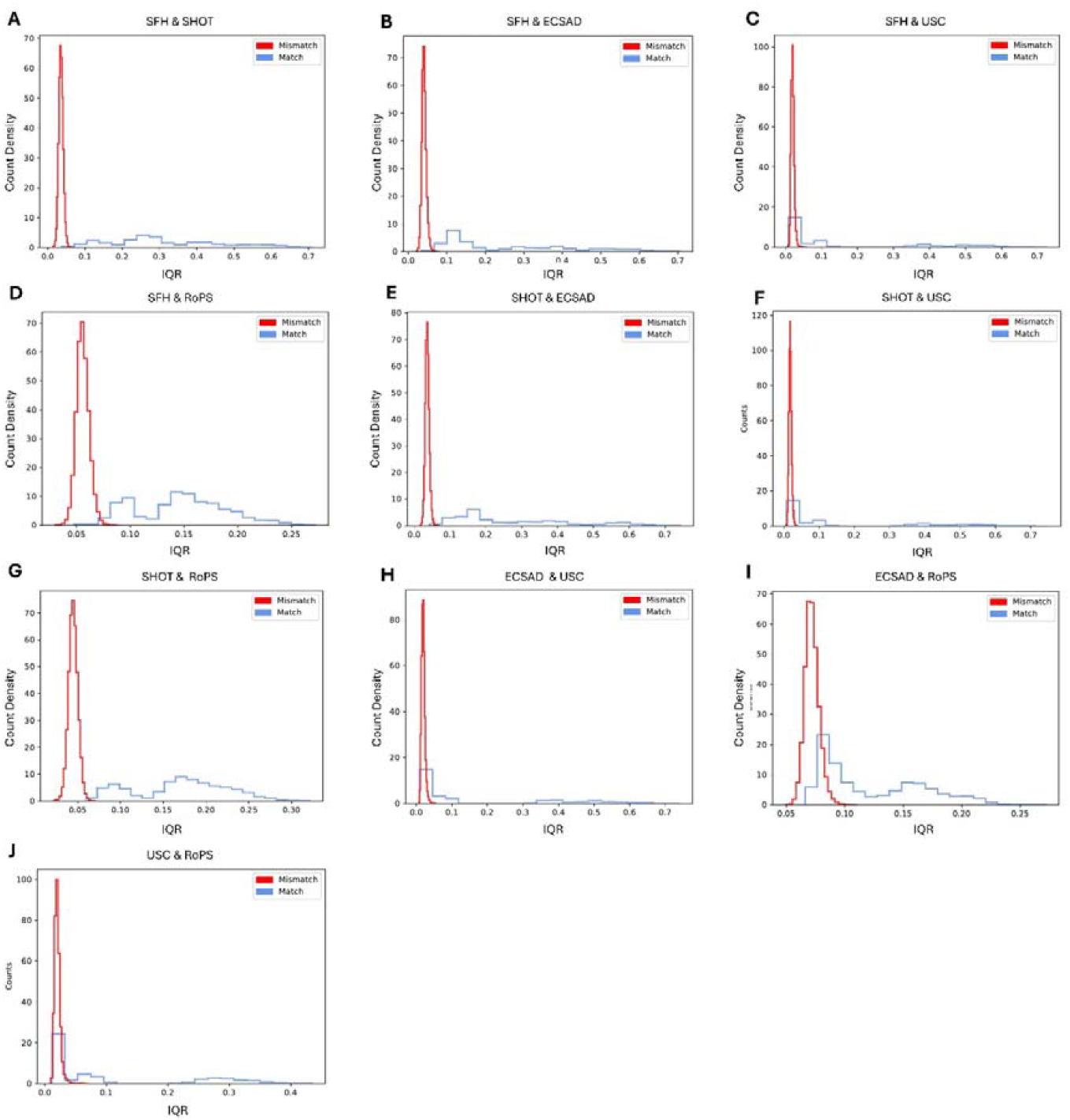
IQR scores of the L2 ratios for each dental scan comparison using combinations of representations. Shown as density histograms. Representation is made using support radius = 2 mm and combinations of representation methods A) SFH & SHOT, B) SFH & ECSAD, C) SFH & USC, D) SFH & RoPS, E) SHOT & ECSAD, F) SHOT & USC, G) SHOT & RoPS, H) ECSAD & USC, I) ECSAD & RoPS, J) USC & RoPS.

## 4 Discussion

This study explores the use of keypoints to distinguish between 3D dental photo scans originating from the same or different human dental arches. We found that the representation method DoC keypoint detection together with SHOT keypoint representation allow for separation of the matches and mismatches in the majority of cases tested in this study. For the present application, combinations of representation techniques did not prove better separation of matches and mismatches and is therefore considered an unnecessary complication of keypoint representation [23]. The representation methods varied in complexity from 30 dimensions to 1980 dimensions [15, 23, 28-31]. A higher complexity causes slower match/mismatch testing but is theoretically able to capture more details about the surface structure. However, in the case of dental surface meshes originating from intraoral 3D photo scans, the higher complexity does not appear to capture the needed variations to unambiguously distinguish between a match and a mismatch. It is therefore not advantageous and results in unnecessary long computation time [15, 17, 22].

Importantly, by using 3D instead of 2D dental imaging data for identification purposes, this study eliminates the variability of shooting angle and image distortion, thus lowering the impact of variance between data acquisition; a known issue within the field [10, 11, 32]. Also, in another subfield of forensic odontology: bitemark analysis, 3D models have been used to lower the distortion caused by 2D imaging [32]. When comparing dentitions to bitemarks, the position of a set of landmarks on the dentition is matched to the placement of landmarks in the bitemark (with or without deformation transformation) [32-34]. This means that the information used for matching is the relative placement of the landmarks. In our study, the relative placement of keypoints is not considered, as all keypoints are compared independently. This ensures that dental position has a minimal effect on the matching score, thus theoretically enhancing robustness of the matching score in the case of tooth loss and changes in individual tooth position. Conversely, the relative placement of keypoints might be able to enhance separability of matches and mismatches, but at the cost of score robustness.

Other methods for dental comparison stemming from 3D dental data exists, though, each with specific pitfalls [12, 15, 16]. The method proposed by Miki *et. al* limits the information used by focussing solely on molars and by reformatting into a 2D image before performing analysis [16]. In the present study we lowered the amount of information by focussing on dental curvatures, thus utilised dental information from all present teeth and kept the 3D aspect in our analysis. The study by Zhou *et. al* shows a very similar logic as the present study, with the setup of dental mesh down sampling, point cloud representation and comparison [12], though, the two studies diverge in specific comparison methodologies. Our study compares the high-resolution surface curvatures in the immediate vicinity of a keypoint, with each keypoint being compared independently. On the contrary, Zhou and colleagues compare relative placement of curvature keypoints using point cloud registration, potentially risking affecting dental matching due to small dental changes, e.g. single tooth loss or change of tooth position [12]. The general similarity in framework underscores the effectiveness and suitability of the overall data processing structure for automated dental comparison. At the same time, the divergence in specific methods employed in each study broadens the exploration of techniques within this emerging area, contributing to a more comprehensive understanding of automated scoring in dental analysis, both aiming at a quantitative measure of dental surface similarity [12].

A possible limitation in our study is the use of SHOT representation when choosing the optimal support radius. By using SHOT in the optimisation, it is possible that bias towards SHOT being the best performing representation method is added to the study. Though, by using a fixed support radius for all representation methods, comparability between methods is guaranteed, but does not eliminate bias. The optimal representation method varies between data types [23]. This study shows that SHOT performs well, but it does not exclude that other representation methods could exceed SHOT performance if variables were optimised differently. If other methods were used for support radius optimisation, the support radius should still fall within the 1mm – 3mm mark, to ensure robustness to teeth positioning.

Further, one could argue that not all information of the dental surface was used for the comparisons in this study. By using keypoints, large parts of the surface information remain unused. The keypoints are focused on curvatures on the dental surface, meaning that most excluded information is from the smooth dental surfaces. The logic for excluding this information is two-fold. Firstly, each full resolution jaw contains several hundred thousand points thus exhausting computational power. By using keypoints, each jaw can be downsized to hundreds to a few thousands keypoints, significantly reducing the computational burden. The time factor is particularly crucial in the event of a disaster, as there is often great pressure from both the authorities, the government, and the public, including worried relatives, to identify the victims as quickly as possible. Secondly, with the lack of curvatures, the smooth dental surfaces are expected to have a high similarity between individuals, which can cause false confidence in an untrue match [15, 17, 22]. Therefore, it is advantageous to exclude the smooth dental surfaces when comparing dental surface scans.

The dentition of a victim will, at least to some degree, change between an AM and a PM scan. Periodontitis, dental caries, erosion, abrasion, natural tooth movement, tooth loss, and adverse disaster conditions are all causes that change the surrounding soft tissue and/or the dentition [2, 12]. Further, different scanners and scanning techniques may cause systematic variation between AM and PM scans [35, 36]. We have tried to simulate dental differences caused by partial dentitions and potential systematic differences caused by scanning an individual twice with the same or even different scanners. The dental mesh comparison in this study replicates a forensic odontology process, as it would be if automated 3D dental scan comparison were used in Disaster Victim Identification (DVI). This study covers a DVI scenario where the AM 3D dental scan would have been recorded by a dentist shortly before the time of death, while the PM 3D dental scan would be conducted by a forensic odontologist immediately following the disaster, thus limiting significant natural dental changes.

A logical next step in this field would be to incorporate a dental dataset containing individuals who have undergone dental scans at multiple time points. This would allow for an evaluation of the performance of the method pipeline when accounting for natural dental changes and the additional noise they introduce.

## 5 Conclusion

In conclusion, the use of keypoint detection, representation, and correspondence to distinguish between intraoral 3D photo scans from the same individual and scans from different individuals is promising in forensic odontology identification. Using Difference of Curvature (DoC) for keypoint detection and Signature of Histograms of OrienTations (SHOT) for keypoint representation, combined with the Inter Quartile Range (IQR) of the L2 ratios of distances between keypoints, was the best tested solution for distinguishing between scans. This approach has the potential to be implemented in the forensic odontology identification process, particularly in cases where the deceased have minimal or no dental work, and where there might be systematic differences between AM and PM dentition data.

## Data and code availability

The research data consists of personal dental data that the authors are not authorized to share. The code used in this study can be found here: https://github.com/AnikaKofodPetersen/Dental-Similarity-Quantification.

## Competing interest

The authors declare that they have no known competing financial interests or personal relationships that could have appeared to influence the work reported in this paper.

## Acknowledgements

We would like to thank AUFF NOVA for funding this study (Grant number: AUFF-E-2021-9-14). Part of the computing for this study was carried out on the GenomeDK cluster. We would like to thank GenomeDK and Aarhus University for providing computational resources and support for this research.

## Notes

### Competing Interest Statement

The authors have declared no competing interest.

### Funding Statement

This study was funded by AUFF Nova (Grant number: AUFF-E-2021-9-14). Part of the computing for this study was carried out on the GenomeDK cluster.

### Author Declarations

The Danish National Committee on Health Research Ethics declared exception of notification for this work

